# Protein Vaccine Demonstrates Less Reactogenicity than mRNA – A Real World Study

**DOI:** 10.1101/2023.05.31.23290594

**Authors:** Matthew Rousculp, Ryan Ziemiecki, Anthony M. Marchese

## Abstract

A prospective, observational study investigated the burden and impact of vaccine reactogenicity in adults from the United States and Canada who received an approved/authorized mRNA COVID-19 vaccine or the Novavax COVID vaccine as a booster dose. As part of the real-world study, reactogenic events captured via patient symptom diaries were compared. Results demonstrated that the overall percentage of Novavax COVID booster recipients experiencing any reactogenicity event (77.6%) was lower than that of mRNA doses (95.9%).

## Introduction

In the United States, decisions regarding COVID-19 vaccine updates are expected to harmonize approaches across platforms and optimize variant-specific immunity. The options will include the widely utilized Pfizer and Moderna mRNA vaccines, and Novavax’s protein-based alternative (NVX-CoV2373). COVID-19 vaccine hesitancy is complex and dynamic in nature, but healthcare providers who are empowered to address their patients’ concerns can make positive impacts in their communities^1-3^. In conjunction with excellent efficacy, mRNA vaccines have been associated with relatively strong reactogenic events which raises concerns that vaccine-induced side effects could threaten future uptake. A survey of healthcare workers indicated the side effects associated with mRNA vaccinations have led to a reduction in their future willingness to receive booster vaccinations^4^. In clinical trials, NVX-CoV2373 has shown similar efficacy, immunogenicity, and safety to mRNA options. Due to its relatively late authorization, little real-world evidence for NVX-CoV2373 is currently available.

## Methods

Local and systemic reactogenic events related to COVID-19 vaccines were compared as part of a prospective, observational study investigating the burden and impact of vaccine reactogenicity. Vaccine recipients from the United States and Canada were recruited through primary care and specialty care clinics. Participants completed a vaccine symptoms diary over six days post-vaccination. The study excluded individuals with confirmed or suspected immunocompromised conditions including active cancer and chronic administration, as well as a history of severe allergic reaction to prior COVID-19 vaccines.

## Results

Eligible working adults ages 18 to 65 years old receiving a single authorized/approved COVID-19 vaccine booster dose were enrolled between July 2022 and March 2023; 827 receiving an mRNA booster (mRNA-1273 [Moderna, Cambridge, MA]; or BNT162b2 [Pfizer, New York, NY]) and 303 receiving NVX-CoV2373 (Novavax, Gaithersburg, MD) booster. The demographic and clinical characteristics of participants were similar, with a median age of 38 years old in both groups **(Table 1)**. Descriptive results indicated that the overall percentage of NVX-CoV2373 booster recipients experiencing any reactogenicity event was 77.6% which is lower than that of mRNA doses, 95.9%. Specific incidences of local and systemic symptoms were captured, and for each, NVX-CoV2373 was found to have a lower percentage of events reported **(Figure 1**). There was a notable difference in the percentage of reported symptoms (mRNA-NVX-CoV2373); injection site tenderness (25.4%), injection site pain (30.4%) muscle pain (30.8%), swelling (22.4%), redness (13%) fatigue (25.7%), malaise (24.9%), headache (18.3%), joint pain (17.5%), fever (12%), nausea/vomiting (3.5%).

**Table 1:** Demographic and Clinical Characteristics of Participants Receiving a Booster Dose

| Characteristic | Individuals, No (%) |  |
| --- | --- | --- |
|  | NVX-CoV2373<br>(n= 303) | mRNA*<br>(n= 827) |
| Age (years) |  |  |
| Mean (SD) | 38.9 (11.8) | 40.1 (13.0) |
| Median | 38 | 38 |
| Minimum, maximum | 18.0, 65.0 | 18.0, 65.0 |
| Age (years) |  |  |
| 18 - 19 years old | 7 (2.3%) | 14 (1.7%) |
| 20 - 29 years old | 70 (23.1%) | 202 (24.4%) |
| 30 - 39 years old | 91 (30.0%) | 214 (25.9%) |
| 40 - 49 years old | 70 (23.1%) | 158 (19.1%) |
| 50 - 59 years old | 52 (17.2%) | 172 (20.8%) |
| 60 - 65 years old | 13 (4.3%) | 67 (8.1%) |
| History of COVID-19 |  |  |
| Yes | 119 (39.3%) | 433 (52.4%) |
| No | 184 (60.7%) | 394 (47.6%) |
| Medical Conditions with High-Risk for COVID-19 |  |  |
| Yes** | 19 (6.3%) | 45 (5.4%) |
| No | 284 (93.7%) | 782 (94.6%) |
| Medical Conditions with High-Risk for COVID-19 |  |  |
| Diabetes | 6 (31.6%) | 21 (46.7%) |
| Hypertension | 7 (36.8%) | 13 (28.9%) |
| Heart disease | 2 (10.5%) | 8 (17.8%) |
| Respiratory conditions | 5 (26.3%) | 11 (24.4%) |
| Other | 5 (26.3%) | 13 (28.9%) |
| Gender |  |  |
| Female | 156 (51.5%) | 469 (56.7%) |
| Male | 142 (46.9%) | 355 (42.9%) |
| Gender fluid | 1 (0.3%) | 0 |
| Nonbinary | 2 (0.7%) | 3 (0.4%) |
| A gender identity not listed | 0 | 0 |
| Prefer not to answer | 2 (0.7%) | 0 |
| Ethnicity |  |  |
| Hispanic | 154 (50.8%) | 207 (25.0%) |
| Non-Hispanic | 149 (49.2%) | 620 (75.0%) |
| Race |  |  |
| African American or Black | 33 (10.9%) | 77 (9.3%) |
| Alaska Native, American Indian, Native American | 2 (0.7%) | 4 (0.5%) |
| Asian | 40 (13.2%) | 189 (22.9%) |
| Middle Eastern or North African | 5 (1.7%) | 21 (2.5%) |
| Native Hawaiian or Pacific Islander | 6 (2.0%) | 75 (9.1%) |
| White | 152 (50.2%) | 278 (33.6%) |
| Other | 65 (21.5%) | 183 (22.1%) |

\*mRNA-1273, or BNT162b2; \*\*If yes, participants selected all specific conditions that applied.

**Figure 1.**
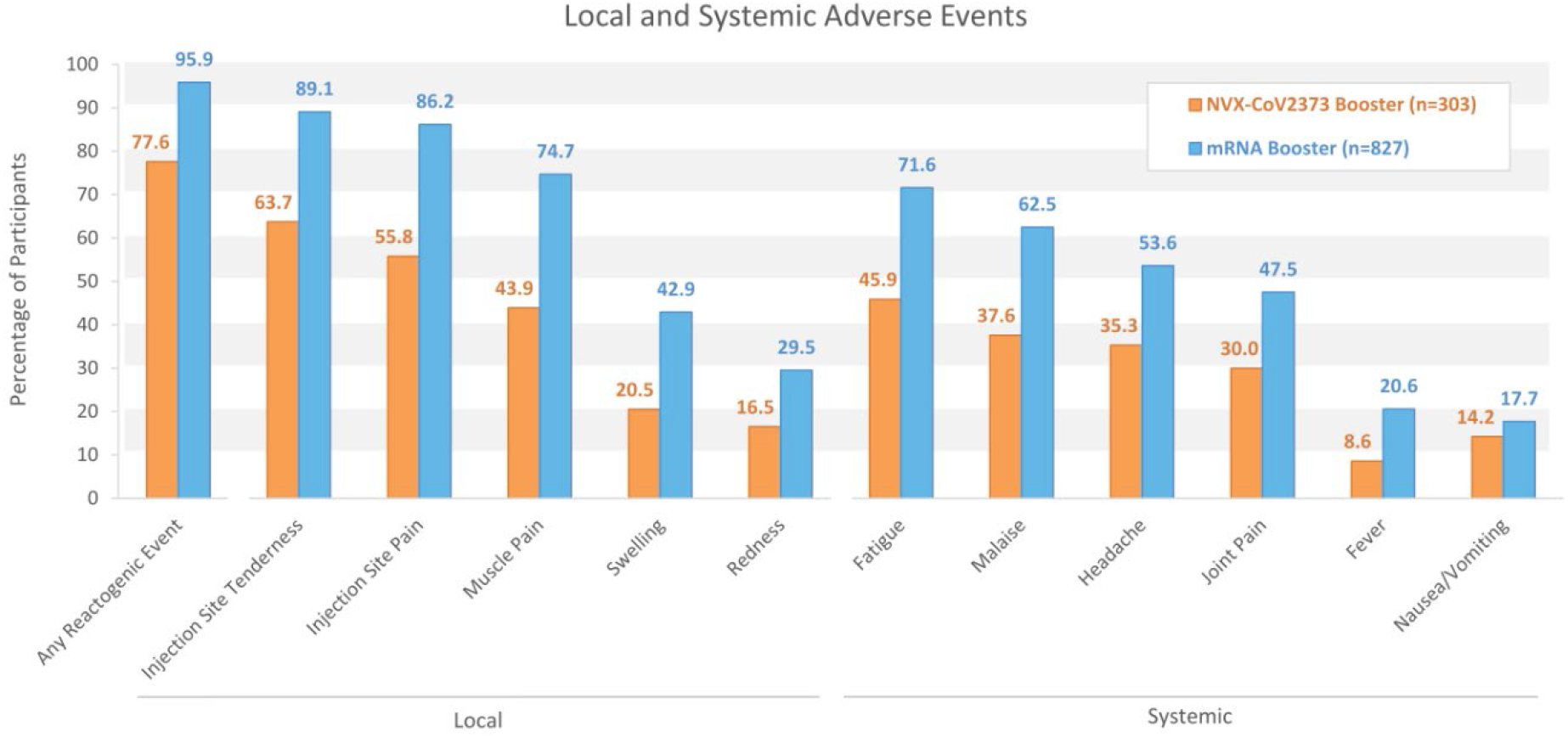
Percentage of Booster Dose Participants Reporting Local and Systemic Adverse Events. Solicited local and systemic adverse events among participants receiving an mRNA booster (blue) or an NVX-CoV2373 booster (orange) with are plotted. Unadjusted results are based on participant diaries collected over 6 days post-vaccination.

## Discussion

These findings support an earlier systematic review and meta-analysis which found mRNA primary series doses to be more reactogenic than other vaccine platforms, including NVX-CoV2373^5^. To combat vaccine hesitancy and promote uptake during future booster campaigns, the ability of vaccine providers to anticipate and respond to concerns in their communities is critical^1^. This information can be influential as healthcare providers engage in conversations with reactogenicity-concerned patients.

Complete analysis of this study, which will investigate the impact of reactogenicity on workplace presentism and absenteeism following primary series and booster doses, will provide additional perspective on the burden of vaccination-associated side effects. This descriptive work is limited by the self-reported nature of patient symptom diaries, and its design as a non-randomized study in US and Canadian adults. Additional work is required to further understand the reactogenicity of different COVID-19 vaccine platforms in different populations. The deployment of the next-generation of COVID-19 vaccines must feature a patient-centric and personalized approach that permits providers the ability to tailor vaccine choice based on the needs and preferences of those they work to protect.

## Data Availability

Requests for data will be considered.

## Article Information

### Author Contributions

Design of study methodology and scientific expertise was provided by MR and AMM. Recruitment, study activities, and data analysis were conducted by RZ. Study supervision and analysis review was led by MR. The manuscript was written, and the figure was prepared by AMM. The manuscript was edited by MR and RZ. All authors revised and approved the final version of the manuscript.

## Acknowledgments

Study design and scientific expertise was provided by Seth Toback and Hadi Beyhaghi of Novavax, Inc. Clinical operations were led by Angela Miller of Novavax, Inc. Research and data analysis support was provided by Kelsey Hollis, Dawn Odom, Ji Min Choi, J. Bradley Layton, Shardul Odak, Laurin Jackson, Val Williams, of RTI Health Solutions. Editorial support was provided by Kelly Cameron, PhD, CMPP of Ashfield MedComms (New York, USA), an Inizio company, supported by Novavax, Inc.

